# The deployment of ProKnow for cloud-based clinical research in radiotherapy

**DOI:** 10.1101/2025.11.24.25340929

**Authors:** Vasiliki Anagnostatou, Martin Knauer, Sebastian H. Maier, Tobias Winderl, Michael Reiner, Reinhard Thasler, Stefanie Corradini, Maximilian Niyazi, Ludwig C. Hinske, Claus Belka, Stephan Schönecker

## Abstract

ProKnow is an archive and restore tool for radiation oncology and imaging data, peer review, distributed contouring, study of metrics and discovery of trends; it represents a common ground for plan analysis and comparison due to an integrated industry standard Dose-Volume Histogram (DVH) engine, which can be used for all patient datasets. The aim of this work is to present a deep and easy-to-use implementation of the Elekta ProKnow DS cloud-based Picture Archiving and Communications in Radiotherapy (RT-PACS) system within our department. Two de-identification workflows of the DICOM data are presented, the first one is accomplished via the ProKnow Dicom Agent (PDA) and the second one involves a trusted third-party service. We can access ProKnow not only through the user interface, but also through the Application Programming Interface (API) with scripts written in the Python language to extract information from the uploaded data, calculate and store metrics as well as upload clinical data. We used ProKnow for a retrospective feasibility study of an isotoxic dose-escalated radiotherapy concept for glioblastoma. Furthermore, to ensure protocol-compliant irradiation planning for the preparation of a prospective dose-escalation trial, we conducted a dummy run with 10 collaborating institutes in Germany. RT-structures were automatically downloaded (via the API) and the Dice Score and Hausdorff Distance were calculated and set as metric in ProKnow. A drawback of the currently implemented de-identification process is that in a subsequent clinical data upload, matching the original and de-identified IDs is not possible. We therefore collaborate with the MeDIC^LMU^(Data Integration Center [DIC]) for development and implementation of an automated de-identification process via a trusted third party service. With this architecture, it will be possible to merge clinical data in local DIC databases with de-identified data in ProKnow at any point in time.

**Author Summary:** Until now, it has been difficult to make scientific comparisons of treatments in radiation oncology because complex dose-volume distributions have to be compared rather than point doses at an isocentre, as is often thought. For the first time, ProKnow software enables the manufacturer-independent comparison of dose-volume distributions in tumour and normal tissue for large cohorts. With the above-described integration, multicentre research with state-of-the-art, secure, cloud-based data flows becomes easy to use.

## Introduction

With the expansion of cloud computing and cloud storage, Picture Archiving and Communications in Radiotherapy (RT-PACS) systems have become available as cloud-based solutions, especially not only for storage but also for viewing and analysis of data for clinical research. ProKnow DS (Elekta, Stockholm, Sweden) is a cloud-based software solution (SaaS) accessed from any device over a web browser and/or an Application Programming Interface (API) written in the Python language for archiving, restoring and managing of radiation oncology data, plan evaluation and “big data analysis”. Software’s build-in tools, such as the calculation of key radiation therapy metrics, the user’s ability to define metrics and set scorecards, organize, analyze and explore cohorts of patients, enable research and outcome analysis anytime from anywhere in the world. Due to the additional ProKnow’s vendor agnostic technology allowing its use with a variety of treatment planning systems render it an attractive tool for big data gathering, benchmarking of planning practices, standardization and clinical excellence. ProKnow was chosen by the NHS England for the above-mentioned capabilities and was deployed in 2019 in 49 radiotherapy providers to enable sharing and learning from radiotherapy plans. It is now populated with more than 26000 patients [1]

Potential in advancements of methodologies and techniques in radiation therapy can be identified in many study areas of radiation therapy in the literature; ProKnow provides a framework for benchmarking of planning practices (international, national, local studies) [1–18] (see also Table 1), testing of segmentation algorithms [19–21], testing of automated treatment planning techniques [22], DVH evaluation [23–24], education training in radiotherapy for physicians and/or medical physicists [25–32], as well as TPS planning conversion validation and archival [33].

**Table 1.** Literature summary on ProKnow usage.

| Year | Tumor Site | Study type | Country | Plans/Patients | Reference |
| --- | --- | --- | --- | --- | --- |
| 2018 | Craniopharyngioma | International | - | 1 | Whitaker et. al. |
| 2020 | Prostate | National | UK | 102 | Taylor et.al. |
| 2020 | Cervix | National | LatAm | 1 | Li et. al. |
| 2022 | Breast | Local | Minnesota,<br>USA | 35 | Morris et. al. |
| 2023 | Adrenal Gland,<br>Pancreas,<br>Oligo metastasis<br>(pelvis),<br>Prostate | International | - | 4 | Bisgaard et. al. |
| 2023 | Pancreas | International | - | 136 | Parikh et. al. |
| 2023 | Breast | International | - | 10 | Tan et. al. |
| 2024 | Craniospinal | National | Turkey | 2 | Senkesen et. al. |
| 2024 | H & N | International | - | 2 | Selek et. al. |
| 2024 | Lung | International | - | 2 | Selek et. al. |
| 2024 | H & N | Local | Minnesota,<br>USA | 136 | Pepin et. al. |
| 2024 | Lung | Local | Newcastle,<br>UK | 102 | Stamp et. al. |
| 2024 | Lung, breast | National | UK | NA | Byrne et. al. |
| 2024 | Prostate, cervix,<br>liver, lymph node | International | - | 5 | Murr et. al. |
| 2024 | Glioblastoma | National | Germany | 10 | Maier et. al. |
| 2024 | H & N | Local | Florida,<br>USA | 20 | Cabrera et. al. |
| 2024 | Glioblastoma | National | Germany | 10 | Bodensohn et.<br>al. |
| 2024 | Prostata | International | - | 5 | Sritharan et. al. |

A literature summary of the international, national and local studies performed with ProKnow

Since the acquisition of the software, we have focused our efforts to build an end-user friendly and individual configurable data pipeline of radiotherapy data (DICOM) from internal clinical databases and locally archived data storages into the ProKnow cloud-based RT-PACS system. The de-identification of the DICOM data, which is a prerequisite for uploading RT-Plans on any cloud-based RT-PACS system in Germany, is part of the dataflow pipeline.

Within our department, we have established ProKnow for internal and nationwide retrospective as well as prospective radiotherapy studies of various tumor sites; we have exploited its capability for remote peer-review, quality assurance in the preparation of a clinical trial (protocol compliance) [13] as well as treatment plan quality evaluation, during a prospective multicenter phase IIa trial [9]. Owing to ProKnow’s API, we have developed scripts in Python, which run automatically on a virtual server following a patient upload to extract information from the uploaded data for calculation and storing of important metrics for plan evaluation and analysis, such as the Equivalent Uniform Dose (EUD), Dice Score and Hausdorff distance, adding value to the clinical decision.

## Materials and Methods

### Pre-configuration of a Dicom Agent service

A dedicated virtual server (Windows Server 2022, CPU 2, RAM 8 GB, partition System: 100 GB, data: 260 GB) had been set up for the installation and configuration of the software tools required for the de-identification and upload of patients’ DICOM data from clinical premises to ProKnow. It is important to note that ProKnow DS does not support any of the DICOM network services (transfer, query/retrieve, workflow management, print management), which is the reason why the workflow for transferring of DICOM data to ProKnow need to include other applications like the ProKnow Dicom Agent (PDA) or the API. The PDA is a local Windows application that supports network transfer of the standard DICOM SOP Classes, and it is recommended to install it on hardware equipped with device encryption to protect data, including protected health information (PHI) and personally identifiable information (PII). In addition to DICOM data transfer, the PDA provides de-identification of selected attributes, as defined, for example, in NEMA PS3 [34].

The PDA is installed on the virtual server and managed by a server administrator who pre-configures a so-called Dicom Agent de-identification service for each project. Since the PDA uses the API directly to upload DICOM files, the user is granted API permission which is encrypted within a .json file that can be download from the user interface. The pre-configuration of a Dicom Agent service includes loading the .json file, loading a user-defined .xml file with the DICOM attributes’ de-identification rules (e.g., “Clear”, “Remove”, etc.), the “Watched” folder destination (original data), the local data storage destination (copies of the original and de-identified data), as well as the data destination in ProKnow (i.e the ProKnow workspace). The original data reaching the “Watched” folder are de-identified based on the .xml file and are uploaded promptly to the ProKnow workspace, while copies of the original and de-identified data can be saved locally on the virtual server.

### Transfer of DICOM data to ProKnow

#### De-identification via the Dicom Agent

The transfer of the DICOM data from the clinical databases (the treatment planning system Monaco and the patient database MOSAIQ) to the “Watched” folder (Process 2, Fig 1), is accomplished using the storescp application of the OFFIS’ open source DICOM Toolkit DCMTK [35], which is installed and run on the virtual server.

**Fig 1.**
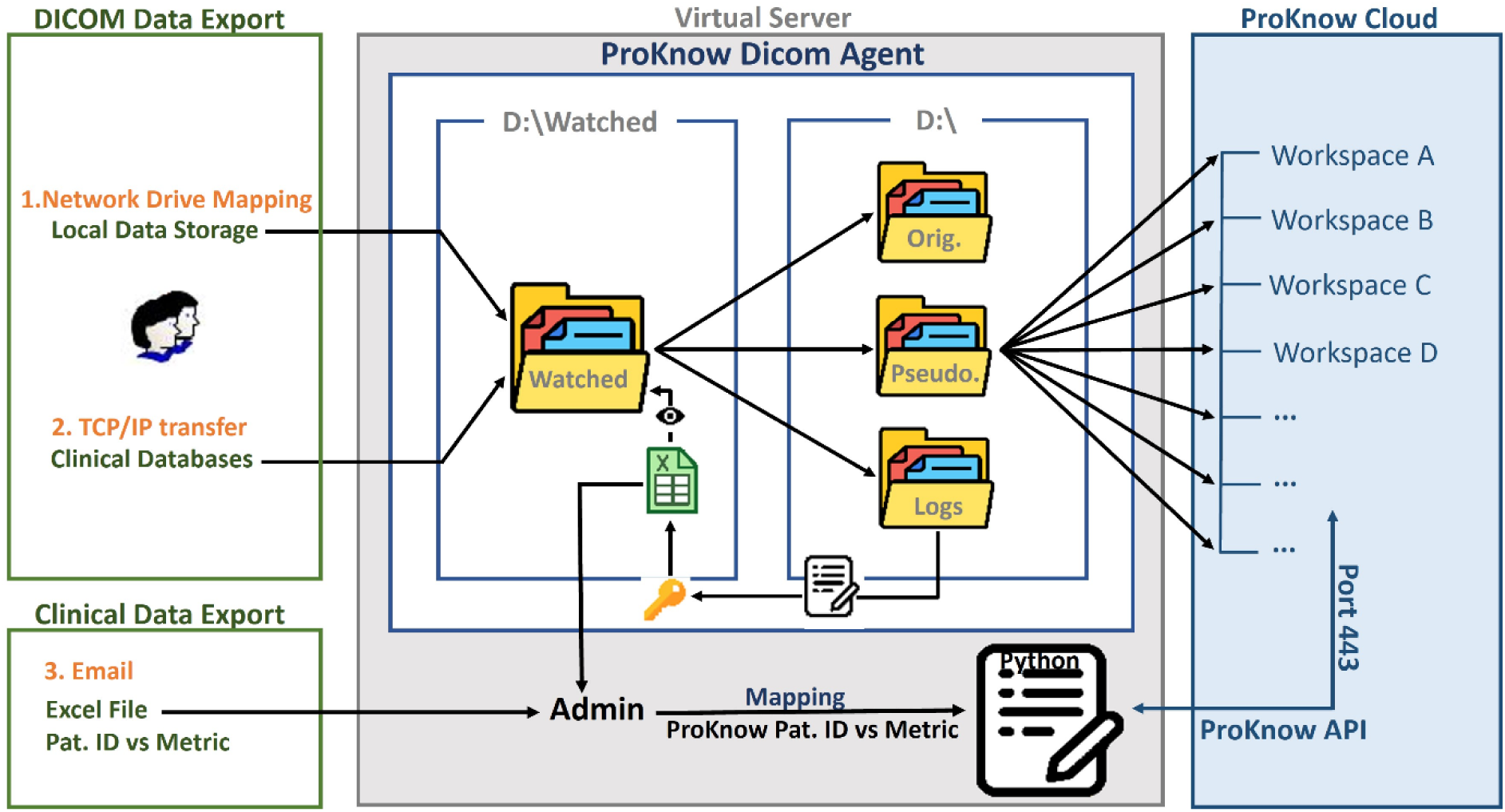
Dicom and Clinical Data de-identification and Upload to ProKnow (via the PDA). Graphical representation of the Dicom and Clinical Data Export routine and de-identification of Clinical DICOM data to ProKnow via the ProKnow Dicom Agent (PDA). Please note that the possibility for a key-list file generation (designated with the key and excel file symbol) visible on the “Watched Folder” can be applicable to prospective trials only.

The aforementioned application implements a Service Class Provider (SCP) for the Storage Service Class, listens on a specific TCP/IP port for incoming association requests from a Storage Service Class User (SCU) and can receive the patients’ DICOM data within a user-defined local folder (in this case the “Watched” folder). On the other side, the clinical databases (SCU user) are configured to export data to the specified IP address and pre-configured TCP/IP port(s) on the virtual server. The transfer of the DICOM data could alternatively be processed directly from the clinical databases to the “Watched” folder, bypassing the use of the DCMTK toolkit; the AE entity and TCP/IP port defined within the PDA de-identification service (SCP user) should be identical to the AE entity and port defined within the export configuration settings of the clinical databases (SCU user).

DICOM data saved within a local folder of the clinical network, can be transferred to the “Watched” folder of the virtual server via network drive mapping; write/read access should be given to the users (e.g., physicians) involved in a project, who can thereafter copy the data to the mapped “Watched” folder (Process 1, Fig 1).

The data reaching the “Watched” folder are automatically handled by the pre-configured PDA service, de-identified and uploaded to a ProKnow workspace, while the generated log .txt files, containing the code list (key-list patient identification), are saved locally on the virtual server. The log files can be sorted and automatically converted to a csv/excel file (using scripting) which can be visible (to the project’s involved users) within the “Watched” folder in case of a prospective study. Following the upload of the de-identified dicom data to ProKnow, the clinical data upload can proceed at any time (Fig 1, bottom), as long as the server administrator maps the original Patient IDs with the ProKnow patient IDs (i.e the patient data should match the de-identified ids). The code list lies on the virtual server and only in prospective studies can be shared to project members. For this reason, the server administrator overtakes the task of the clinical data upload either via the ProKnow User Interface (https) using an excel file of a specified format (with the ProKnow Patient ID and custom metrics) or via the API using a Python script for the assignment and storage of the given metrics in ProKnow.

#### De-identification via a trusted third party

As an alternative to the PDA de-identification service, we are currently developing an automated de-identification and pseudonymization workflow in collaboration with the MeDIC^LMU^ (Data Integration Center) of the LMU University Hospital for the development and implementation of a de-identification process via a trusted third party service. Within this workflow the DICOM data can be transferred from the clinical databases (the treatment planning system Monaco and the patient database MOSAIQ) to a local folder on the virtual server following the processes explained in previous sub-section. As shown in Fig 2, the original patient’s DICOM data are automatically zipped after reaching the local folder on the virtual server (via an automated Python script “watching” the input folder), and sent to the DIC SFTP Server for de-identification. The retrieval of the zipped output data to a “Watched” folder of the virtual sever (as well as unzipping) is undertaken by the automated Python script and is completed within 2-3 minutes. At this stage the Dicom Agent de-identification service is facilitated and can be pre-configured to upload the DICOM data to ProKnow without the use of a .xml de-identification file, since these are already de-identified data.

**Fig 2.**
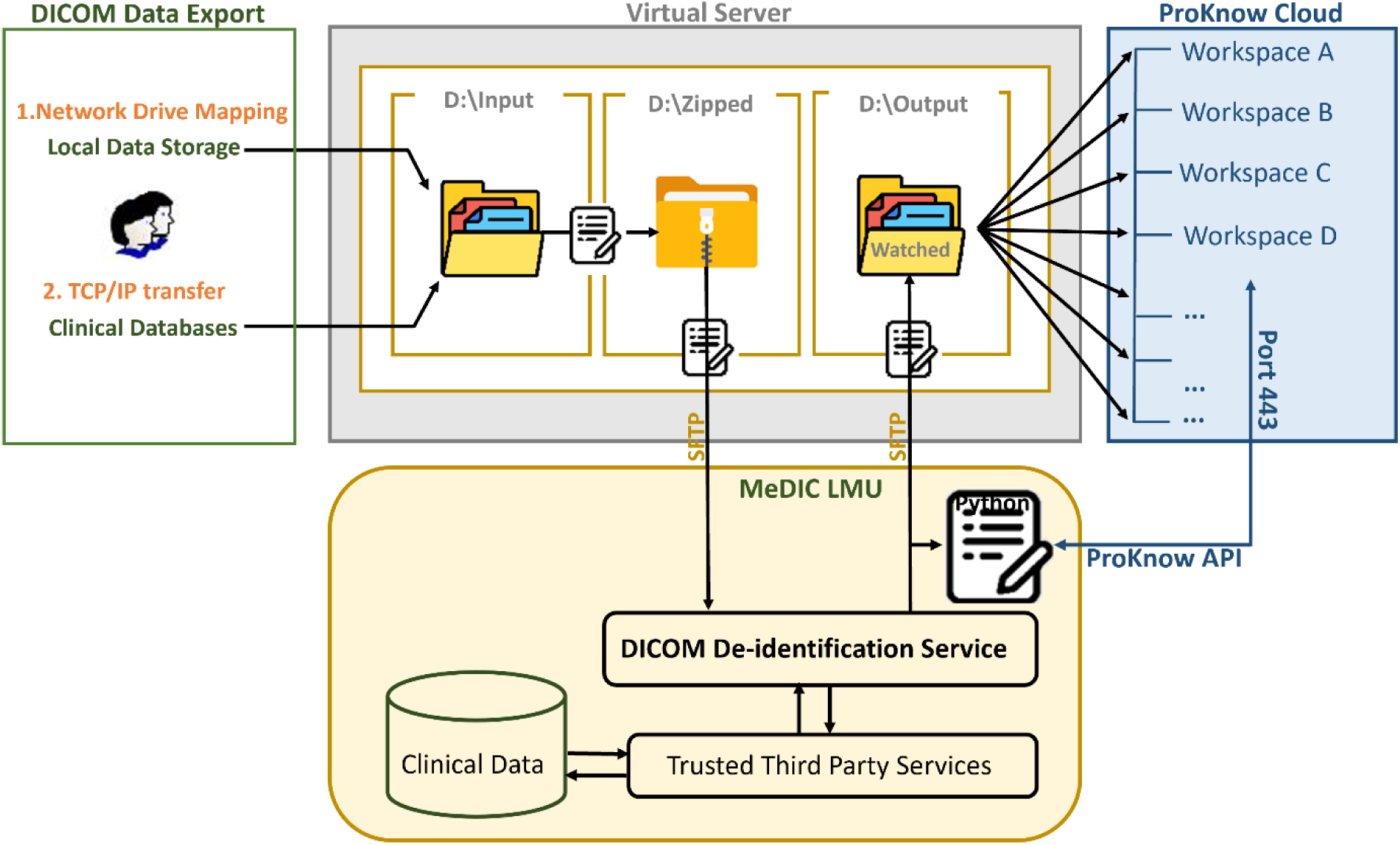
Dicom and Clinical Data de-identification and Upload to ProKnow via the MeDIC^LMU^. Graphical representation of the Dicom and Clinical Data Export routine and de-identification of Clinical DICOM data to ProKnow via the MeDIC^LMU^ (Data Integration Center) and upload via the ProKnow Dicom Agent (PDA).

The MeDIC service results in retaining a code list in a trusted third party service (not known to any project member), that allows longitudinal data enrichment. Clinical Data retrieval could be done in parallel and mapped directly to the de-identified IDs. The data can be directly uploaded from MeDIC to ProKnow, through the ProKnow API, given the fact that DIC will be granted API rights; Nevertheless, a file upload (via https) could also be possible for physicians or other project members, which could be made available into the “Output” folder.

### ProKnow data extraction

The ProKnow API can also be used for extracting information from the uploaded data (such as DVH data) for the calculation of important parameters, as well as storing of these parameters as custom metrics in ProKnow. The API requests in ProKnow (executed through Port 443) require the installation of the Python programming language [36], the installation of the “proknow” python library from PyPI [37], as well as a development environment for programming in Python. For the structure geometric similarity comparison in ProKnow and calculation of the Dice similarity coefficient (DSC) and Hausdorff Distance (HD, and 95th percentile HD95), the open-source software Plastimatch [38] was installed and initiated via a Python script.

## Results

### Implementation - EUD Calculation

A script (attached, S1 Python Script) written in Python running on the virtual server was used to access the information from the DVH graphs for the calculation of the Equivalent Uniform Dose (EUD, introduced by Niemierko et al. [39]) of the brain and storing of the metric in ProKnow (via the ProKnow API), during a retrospective feasibility study of an isotoxic dose-escalated radiotherapy concept for glioblastoma.

### Implementation - DICE similarity coefficient and Hausdorff distance calculation

For a comparison of the geometric similarity between structures a Python Script (attached, S2 Python Script) was developed and run on the server for downloading the respective RT-Structures, initiating the calculation of the Dice similarity coefficient (DSC) and Hausdorff Distance (HD, and 95th percentile HD95), and storing the similarity metric computations in ProKnow. The similarity metric computations are then calculated using Plastimatch, while the downloading of the structures and storing of the metrics are performed via the ProKnow API. The script was used for the similarity comparison between the structures of the main study center and different study sites during the preparation of a prospective dose-escalation trial.

### Use cases

ProKnow was used for the assessment of the dosimetric feasibility of an isotoxic dose-escalated radiotherapy concept for glioblastoma [13], prior to its implementation in the PRIDE trial (“Protective VEGF Inhibition for Isotoxic Dose Escalation in Glioblastoma”, NCT05871021; NOA-28; ARO-2024-01; AG-NRO-06). For the dosimetric analysis of the study, 10 patients, who had previously undergone treatment for glioblastoma, were selected from the internal database MOSAIQ for the comparison and analysis of the standard treatment approach and experimental treatment protocol which was going to be utilized in the PRIDE trial. The images, registrations, structure sets, and dose distributions of the reference and experimental plan (generated in Monaco TPS), were de-identified and uploaded in ProKnow (via the PDA, as described above). The review of the dose values of the target volumes for both plans, the dose exposure of organs at risk (OAR) as well as the dose-volume histograms (DVH graphs) were made available by ProKnow’s dosimetric tools (Fig 3).

**Fig 3.**
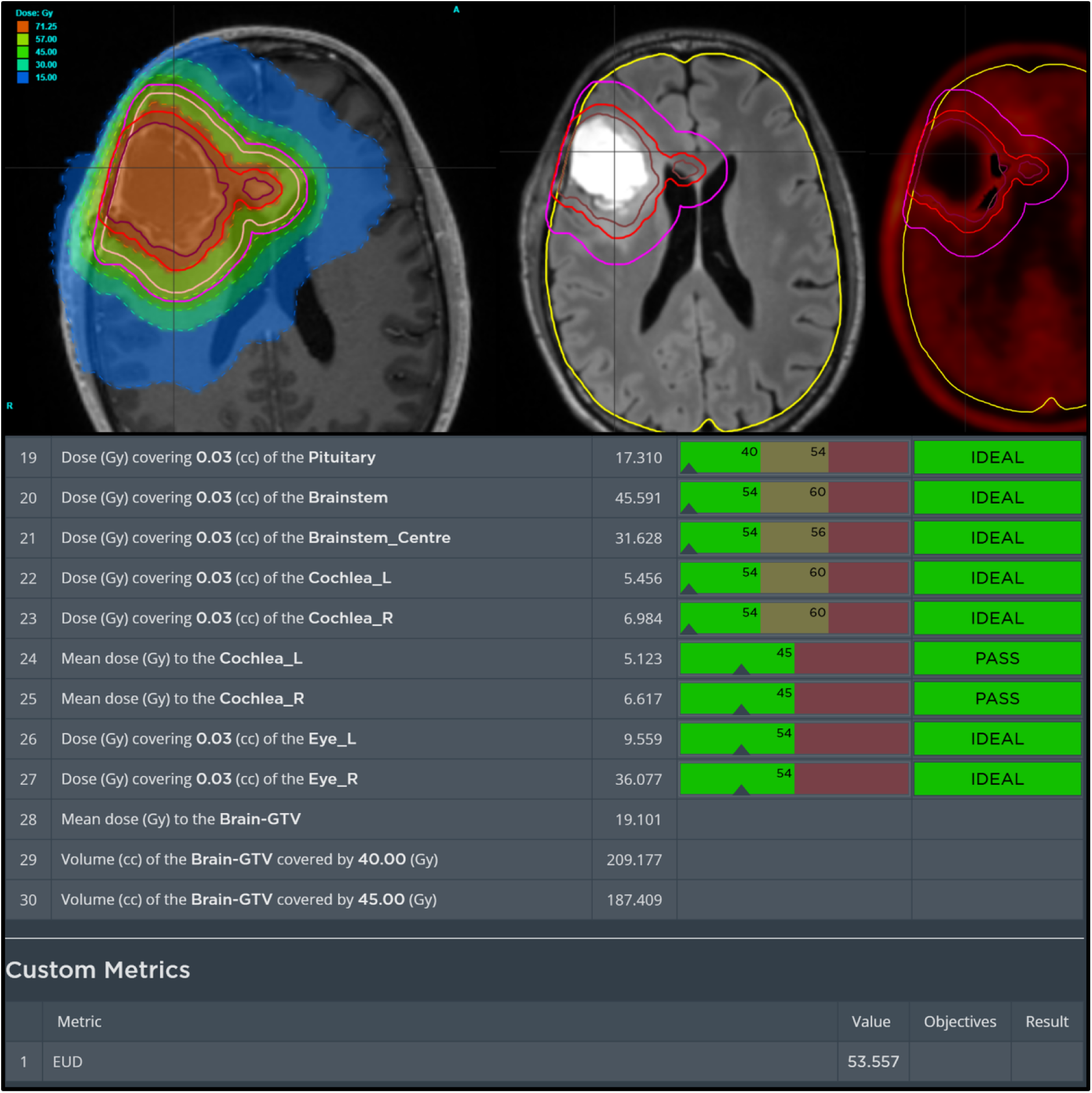
Dosimetric tools in ProKnow. Examples Views made visible for peer-review by the main investigator (PRIDE Dummy Run study [9]), showing MR and PET fusions overlayed over dose and structures in the upper part and a scorecard for a quick overview as well as the automatically calculated EUD.

Based on the prescription and constraints for the study, scorecards were created in ProKnow enabling the assessment and comparison of plan quality. The EUD (see section ’Implementation – EUD Calculation’) was used for the comparison of the reference and experimental treatment plans regarding the assessment of the risk of a radiation treatment-related toxicity, such as radiation necrosis.

Following the demonstration of the above-mentioned treatment concept, ProKnow was also used for the preparation and quality management assurance of the PRIDE trial (Fig 3). To ensure protocol-compliant conduct of irradiation planning with all participating study centers, one benchmarking case (dummy run) from the test cohort from the feasibility study was sent to the collaborating centers. Each study site prepared a radiation plan for the standard as well as a dose escalated plan (according to the study’s manual), including the relevant OAR. The de-identified DICOM plans and dose files were returned to the main study center (via databox – due to data security clearance delays in participating centers), which were uploaded to ProKnow via the PDA (Process 1, Fig 1). Similar to the feasibility study, the EUD of the brain (for both standard and experimental plan) was calculated for each dummy run and set as a custom metric in ProKnow via the ProKnow API. For an exact comparison of the geometric similarity between the structures of the main study center and different study sites, the Dice similarity coefficient and Hausdorff distance were calculated as described in section ’Implementation – DICE similarity coefficient and Hausdorff distance calculation’.. A review process based on the specified parameters was performed by three radiation oncology specialists for all submitted plans, including examining registration, plan quality, and adherence to protocol.

In the prospective PRIDE phase II trial, the first three plans of each center must be approved by the leading study center prior to therapy initiation, which is inherently time-critical in order to avoid delays in treatment. To support this process, the Python script calculating the EUD continuously monitors the ProKnow workspace and automatically initiates the calculation whenever a plan is uploaded. Thanks to the drive-mapping functionality, the upload of a plan to ProKnow can be made from any individual granted access to the “Watched” folder, which automatically triggers the calculation of the EUD without the intervention of the server administrator. The script runs via the task scheduler on the virtual server every 5 minutes, ensuring that the EUD custom metric is generated automatically following a plan upload, thus allowing the main investigator to peer-review the case remotely, assess the plan quality and protocol-compliance, and grant permission for irradiation with the dose-escalated concept.

## Discussion

Uploading of clinical data is a strong feature of ProKnow, allowing to study trends and compare treatment outcomes by observing the correlations between clinical data and/or dosimetric parameters for a cohort of patients (via histograms or scatter plots); it requires, however, the processing of the non-standardized internal files containing the custom metrics as well as the correlation of the original Patient ID with the de-identified ProKnow ID before proceeding to the upload. The clinical data need first to be gathered from a physician or medical student out of many different clinical databases and manually typed into a file (usually an excel file), which is a time-consuming and prone to error procedure. Nonetheless, the data follow a non-standardized format, e.g., a long list of unnecessary for ProKnow PHP data, clinical data listed under the Patient IDs (in subsequent rows) or next to the Patient IDs (in subsequent columns), etc. Such format generates the laborious task of having to manually tailor the file to the ProKnow desired format (upload via User Interface) or update the Python script each time (upload via the API) to perform the correct assignment of the custom metrics to the de-identified patients in ProKnow (Fig 1, bottom). To overcome this issue completely, the solution would be to use merged clinical data in a standardized data format with already de-identified Patient IDs. The Medical Informatic Initiative – Germany – has already established DIC centers in various hospital sites in Germany who are responsible for the data integration, data quality and data protection [40]. For making such solution realizable we have started discussions with the MeDIC^LMU^ (Data Integration Center) of the LMU University Hospital, to undertake the tasks of merging the clinical data in local DIC databases, as well as the development and implementation of an automated de-identification and pseudonymization process via a trusted third-party service (E-PACS), depicted in Fig 2. The file containing the merged clinical data could be retrieved from the SFTP DIC Server via an automatic script or could be sent directly from DIC Server to ProKnow via the API. Within this standardized clinical data file, the de-identified Patient IDs match the ProKnow IDs (the uploaded DICOM data), hence no tailoring or processing of the given file would be necessary for the upload. Clinical or DICOM data can be added to the same patients at any point in time, since PDA keeps a Patient ID database ensuring uploads to the correct patient(s). The establishment of such workflow for the integration of radiotherapy data (DICOM RT) with clinical data and upload to ProKnow, aims to facilitate the discovery of trends and longitudinal outcomes analysis correlated with individual treatment plan data. ProKnow, being an online big-data cloud-based discovery software can be used for the examination of this data between the sites for example in the Bavarian Cancer Research Center (BZKF), opening new opportunities for research and improved healthcare in radiotherapy.

After an extended implementation phase, the advantages of deep implementation include shared network project folders for physicians, customisable access options, and project-specific semi-automated de-identification and data processing in the background.

## Conclusions

We present an easy-to-use implementation of a highly efficient workflow solution that integrates ProKnow into the clinical environment, allowing for project-specific de-identification and post-processing. Once this has been achieved, the way is open for a larger-scale introduction into the radio-oncology study landscape. The possibility of quick, cloud-based processing of DICOM RT data for time-dependent multicentre studies, as well as semi-automated quality control even before the start of radiotherapy treatments during oncological studies, should enable us to take radiotherapy treatment and homogeneity in future studies to a new level.

## Data Availability

The python scripts (provided as supporting files S1 Python Script and S2 Python Script), can be used for the calculation of the metrics presented in the Results section of this manuscript and serve as a guide for similar operations using the ProKnow API.

## Acknowledgments

We would like to thank Rahul Kashyap (Applications Specialist ProKnow, Elekta) and the ProKnow Community Team for their support.

## Supporting information

**S1 Python script. EUD Calculation.** Python script using the ProKnow API to access the DVH graphs for the calculation of the Equivalent Uniform Dose (EUD, [39]) of the brain and store the metric in ProKnow. Please note that the script is saved in a plain text document; to run the script it should be renamed with a .py extension.

**S2 Python script. DICE similarity coefficient and Hausdorff distance calculation.** Python script using the ProKnow API to download the dummy-run RT-Structures, initiate the calculation of the Dice similarity coefficient (DSC) and Hausdorff Distance (HD, and 95th percentile HD95), and store the similarity metric computations in ProKnow. Please note that the script is saved in a plain text document; to run the script it should be renamed with a .py extension

## References

1. Byrne J, Warren S, Walker C. 1661: National benchmarking of lung SABR and breast radiotherapy treatment plans using cloud-based ProKnow. Radiotherapy and Oncology. 2024;194:S4833–S7.

2. Senkesen O, Tezcanli E, Alkaya F, Ispir B, Catli S, Yesil A, et al. Current practices of craniospinal irradiation techniques in Turkey: a comprehensive dosimetric analysis. Radiat Oncol. 2024;19(1):49.

3. Selek U, Hu KS, Tatli H, Palta JR, Jhingran A. Global Interobserver Variations of Prescription and Plan Evaluation within Heterogeneous Clinical Target Volume (CTV) Delineations among International Experts in Head and Neck Cancers. International Journal of Radiation Oncology*Biology*Physics. 2024;120(2, Supplement):e788.

4. Selek U, Hu KS, Tatli H, Palta JR, Jhingran A. Global Interobserver Variations of Clinical Target Volume (CTV) Delineation among International Experts in Given Standard Gross Tumor Volume (GTV) and Organs at Risk (OAR) Volumes in Lung Cancer. International Journal of Radiation Oncology*Biology*Physics. 2024;120(2, Supplement):e788.

5. Pepin MD, Anaya S, Garces YI, Hosfield E, Lester SC, Ma DJ, et al. Radiation Dose Sensitivity of Subregions of the Larynx to Patient-Reported Swallowing Outcomes. Advances in Radiation Oncology. 2024;9(5):101458.

6. Stamp A, Pickles R, Warren S, Smith E, McDonald FE, Hassani A, et al. 143 Using ProKnow to explore the incidence of radiation pneumonitis in relation to GTV size in Stage III Non-Small Cell Lung Cancer (NSCLC) concurrent Chemoradiotherapy (cCRT) patients and its impact on survival. A Pilot Study. Lung Cancer. 2024;190:107704.

7. Taylor T, Richmond N. A UK wide study of current prostate planning practice. Br J Radiol. 2020;93(1111):20200142.

8. Murr M, Bernchou U, Bubula-Rehm E, Ruschin M, Sadeghi P, Voet P, et al. A multi-institutional comparison of retrospective deformable dose accumulation for online adaptive magnetic resonance-guided radiotherapy. Phys Imaging Radiat Oncol. 2024;30:100588.

9. Maier SH, Schonecker S, Anagnostatou V, Garny S, Nitschmann A, Fleischmann DF, et al. Dummy run for planning of isotoxic dose-escalated radiation therapy for glioblastoma used in the PRIDE trial (NOA-28; ARO-2024-01; AG-NRO-06). Clin Transl Radiat Oncol. 2024;47:100790.

10. Cabrera J, Erhart K, Kelly P, Zeidan OA, Swanick C, Rineer J, et al. Defining the role of intensity modulation in electron conformal therapy for the treatment of head and neck cancer. Medical Dosimetry. 2024;49(4):359–62.

11. Li B, Engwo A, Perez T, MacDuffie E, Hao J, Trejo JM, et al. Variability of Current Clinical Practices for Locally Advanced Cervical Cancer through Assessment of Contouring, Prescription, and IMRT/VMAT Planning Abilities. International Journal of Radiation Oncology, Biology, Physics. 2020;108(3):e423–e4.

12. Bisgaard ALH, Keesman R, van Lier ALHMW, Coolens C, van Houdt PJ, Tree A, et al. Recommendations for improved reproducibility of ADC derivation on behalf of the Elekta MRI-linac consortium image analysis working group. Radiotherapy and Oncology. 2023;186:109803.

13. Bodensohn R, Fleischmann DF, Maier SH, Anagnostatou V, Garny S, Nitschmann A, et al. Dosimetric feasibility analysis and presentation of an isotoxic dose-escalated radiation therapy concept for glioblastoma used in the PRIDE trial (NOA-28; ARO-2022-12). Clin Transl Radiat Oncol. 2024;45:100706.

14. Sritharan K, Akhiat H, Cahill D, Choi S, Choudhury A, Chung P, et al. Development of Prostate Bed Delineation Consensus Guidelines for Magnetic Resonance Image-Guided Radiotherapy and Assessment of Its Effect on Interobserver Variability. Int J Radiat Oncol Biol Phys. 2024;118(2):378–89.

15. Parikh PJ, Lee P, Low DA, Kim J, Mittauer KE, Bassetti MF, et al. A Multi-Institutional Phase 2 Trial of Ablative 5-Fraction Stereotactic Magnetic Resonance-Guided On-Table Adaptive Radiation Therapy for Borderline Resectable and Locally Advanced Pancreatic Cancer. Int J Radiat Oncol Biol Phys. 2023;117(4):799–808.

16. Proceedings to the 7th Annual Conference of the Particle Therapy Cooperative Group North America (PTCOG-NA). International Journal of Particle Therapy. 2022;8(4):82–122.

17. Proceedings to the 4th Annual Conference of the Particle Therapy Cooperative Group North America (PTCOG-NA). International Journal of Particle Therapy. 2018;4(4):47–109.

18. Tan IZ, Mitchell A, McNair H, Dunlop A, Herbert T, Nartey J, et al. A Multicenter Study of Clinical to Planning Target Volume Margins for Adjuvant Partial Breast Irradiation Delivered on the 1.5T MR-Linear Accelerator. International Journal of Radiation Oncology*Biology*Physics. 2023;117(2, Supplement):e725.

19. Gibbons E, Hoffmann M, Westhuyzen J, Hodgson A, Chick B, Last A. Clinical evaluation of deep learning and atlas-based auto-segmentation for critical organs at risk in radiation therapy. J Med Radiat Sci. 2023;70 Suppl 2(Suppl 2):15–25.

20. Constantinou AD, Hoole A, Wong DC, Sagoo GS, Alvarez-Valle J, Takeda K, et al. OSAIRIS: Lessons Learned From the Hospital-Based Implementation and Evaluation of an Open-Source Deep-Learning Model for Radiotherapy Image Segmentation. Clin Oncol (R Coll Radiol). 2024;37:103660.

21. Costea M, Zlate A, Serre AA, Racadot S, Baudier T, Chabaud S, et al. Evaluation of different algorithms for automatic segmentation of head-and-neck lymph nodes on CT images. Radiother Oncol. 2023;188:109870.

22. Ceylan C, Kandemir R, Şahin IŞ, Söke Ö, Starbuck W, Can S, et al. PP20.14 SCRIPTING AUTOPLANNING APPROACH FOR EARLY-STAGE PROSTATE CANCERVMAT PLANNING UNDER RTOG 415 RECOMMENDATIONS IN MONACO. Physica Medica. 2024;125:103748.

23. Penoncello GP, Voss MM, Gao Y, Sensoy L, Cao M, Pepin MD, et al. Multicenter Multivendor Evaluation of Dose Volume Histogram Creation Consistencies for 8 Commercial Radiation Therapy Dosimetric Systems. Pract Radiat Oncol. 2024;14(3):e236–e48.

24. Walker LS, Byrne JP. Clinical impact of DVH uncertainties. Medical Dosimetry. 2024.

25. Shanbhag N, Sumaida A, Saleh M. Achieving Exceptional Cochlea Delineation in Radiotherapy Scans: The Impact of Optimal Window Width and Level Settings. Cureus. 2023;15.

26. Naimi Z, Bohli M, Ben Rejeb M, Amor RB, Ghorbel L, Yahyaoui S, et al. Enhancing Childhood Cancer Care In African Low-Middle Income Countries: A Tunisian Pilot Experience. International Journal of Radiation Oncology*Biology*Physics. 2022;114(5):1073.

27. Dizendorf E, Chopra S, Mittal P, Gupta A, Nout R, Sturdza A, et al. Gynecological brachytherapy hybrid training: The Tata Memorial Centre and BrachyAcademy experience. Brachytherapy. 2024;23(6):648–59.

28. Walker K, Chavis Y, Leach D, McLaughlin C. Cloud-based Self-directed Contouring Modules in Radiation Oncology Residency: Interim Results. International Journal of Radiation Oncology*Biology*Physics. 2024;119(4):e15.

29. Roumeliotis M, Morrison H, Conroy L, Becker N, Logie N, Grendarova P, et al. Competency-Based Medical Education in Radiation Therapy Treatment Planning. Practical Radiation Oncology. 2022;12(3):e232–e8.

30. Dizendorf E, Miriyala R, Sreelakshmi KK, Tatli H, Mahantshetty U. PO 0602: Implementation of an Online Contouring Workshop for CT-Based Image-Guided Adaptive Brachytherapy in Cervical Cancer: The BrachyAcademy Experience. Brachytherapy. 2024;23(6, Supplement):S149–S50.

31. Carlson CM, Zhu H, Dempsey C, Shulman A, Biancia CD, Keiper T, et al. Large-Scale Remote Training for Medical Physicists to improve Intensity-Modulated Radiation Therapy/Volumetric Modulated Arc Therapy in Low and Middle-Income Countries. International Journal of Radiation Oncology*Biology*Physics. 2023;116(3):e3.

32. Johnson PB, Schubert L, Kim GG, Faught J, Buckey C, Conroy L, et al. AAPM WGPE report 394: Simulated error training for the physics plan and chart review. Med Phys. 2024;51(5):3165–72.

33. Rusu SD, Atienza C, Smith BR, St-Aubin JJ, Hyer DE. Validating a framework for Pinnacle plan conversion and archival. J Appl Clin Med Phys. 2025;26(9):e70222.

34. Committee DS. NEMA PS3 2025 [Available from: https://dicom.nema.org/medical/dicom/current/output/html/part01.html.

35. Marco E, Joerg R, Thomas W, Andrew JH, Andreas B, Peter J, editors. Ten years of medical imaging standardization and prototypical implementation: the DICOM standard and the OFFIS DICOM toolkit (DCMTK). ProcSPIE; 2004.

36. Foundation PS. Python Programming Language 2001-2025 [Available from: https://www.python.org.

37. Foundation PS. Python Package Index (PyPI) 2025 [Available from: https://pypi.org.

38. Sharp G, Li R, Wolfgang J, Chen G, Peroni M, Spadea M, et al. PLASTIMATCH– AN OPEN SOURCE SOFTWARE SUITE FOR RADIOTHERAPY IMAGE PROCESSING 2010.

39. Niemierko A. Reporting and analyzing dose distributions: A concept of equivalent uniform dose. Medical Physics. 1997;24(1):103–10.

40. Germany MII. Data integration centres [Available from: https://www.medizininformatik-initiative.de/en/consortia/data-integration-centres.

